# Integrated cardiometabolic and nutritional risk profiling identifies pregnancy loss as a marker of systemic metabolic vulnerability

**DOI:** 10.64898/2026.06.04.26354910

**Authors:** Tanya Agarwal, Janaki Ramya Namburu, Priyadarshini Kachroo

**Author notes:** Corresponding author: Department of Health Informatics, Rutgers School of Health Professions, Rutgers-The State University of New Jersey, 65 Bergen Street, Newark, New Jersey 07107, United States.

## Abstract

**Background:** Pregnancy loss has important implications for women’s health. Although maternal age is a well-established risk factor, the contribution of routinely measured cardiometabolic and behavioral markers at population-scale remains incompletely characterized.

**Objective:** To examine associations between cardiometabolic, nutritional, and behavioral risk markers and pregnancy loss among U.S. women of reproductive age.

**Methods:** We conducted a cross-sectional analysis of 4,842 U.S. women aged 20–44 years with ≥1 pregnancy using the National Health and Nutrition Examination Survey data (2013–2023). Pregnancy loss was defined as ≥1 prior miscarriages. Exposures included body mass index, smoking exposure (cotinine), lipid biomarkers, vitamin D and folate, and a composite cardiometabolic–nutritional risk score. Survey-weighted logistic regression estimated adjusted odds ratios (aORs) and 95% confidence intervals, with bootstrap resampling for predictor robustness.

**Results:** The weighted prevalence of pregnancy loss was 23%. Higher odds of pregnancy loss were associated with increasing age (aOR per year=1.02; 95% CI: 1.00–1.04), Non-Hispanic Black race (aOR=1.32; 95% CI: 1.00–1.74), overweight (aOR=1.56; 95% CI: 1.16–2.11), obesity (aOR=2.06; 95% CI: 1.39–3.05), and smoking (aOR=1.58; 95% CI: 1.19–2.10).

Adverse lipid profiles, particularly elevated triglycerides (aOR=1.83; 95% CI: 1.16–2.90) and high low-density lipoprotein (aOR=2.97; 95% CI: 1.45–6.61), were independently associated with pregnancy loss. Vitamin D/folate were not stable predictors. Higher composite cardiometabolic–nutritional risk scores were observed among women with pregnancy loss (P=0.026).

**Conclusion:** Pregnancy loss clustered with adverse cardiometabolic and behavioral risk markers in a nationally representative population. These findings highlight pregnancy loss as a marker of broader metabolic vulnerability supporting the need for longitudinal studies and cardiometabolic profiling to inform preconception care and risk stratification.

## Introduction

Adverse pregnancy outcomes, regardless of the underlying cause, can have lasting psychological and medical consequences for women and families [1]. Recent national estimates suggest that pregnancy loss remains common; Forrest et al. reported that roughly one in five pregnancies among U.S. reproductive-age women ended in loss between 2000 and 2018 [2]. Although advancing maternal age is a significant determinant, several modifiable factors, including cardiometabolic status, nutritional deficiencies, and behavioral exposures, also influence early gestational viability [1]. A growing body of research continues to accumulate for the roles of excess adiposity, tobacco exposure, and metabolic-nutritional disturbances in pregnancy loss.

High body mass index (BMI) before conception or in early pregnancy has been consistently associated with an elevated risk of miscarriage in population-based studies [3]. Tobacco exposure shows a similar pattern, with studies demonstrating a dose–response increase in risk among active smokers; biomarker-based measures such as serum cotinine provide objective evidence reinforcing this association [4].

Micronutrient and metabolic profiles are also plausible contributors to early pregnancy loss. Vitamin D supports immune regulation and placentation, and its deficiency has been linked to a higher risk of miscarriage in pooled analyses [5]. Folate is essential for early embryogenesis, and although supplementation reduces the incidence of neural tube defects, studies on folate status and miscarriage have yielded inconsistent results [6]. Associations between lipid fractions and early pregnancy loss are still emerging, but dyslipidaemia may reflect systemic metabolic and inflammatory pathways relevant to implantation and placental development. Their established relevance to long-term cardiovascular risk further supports evaluating lipid profiles during these critical windows, alongside traditional risk factors, both to improve understanding of pregnancy loss and to inform broader preventive health strategies [7].

The National Health and Nutrition Examination Survey (NHANES) provides nationally representative data, integrating standardised interviews, physical examinations, and laboratory assays within a complex, multistage sampling design [8]. Using NHANES, the present study examines behavioral (smoking), anthropometric (BMI), and nutritional–cardiometabolic indicators (vitamin D, folate, and lipid fractions) within a single analytic framework, adjusting for key sociodemographic confounders, to estimate their associations with pregnancy loss among U.S. women of reproductive age.

## Materials and methods

### Study design, data source, and study population

This study employed a cross-sectional design, utilizing publicly available data from the U.S. National Health and Nutrition Examination Survey (NHANES), provided by the Centers for Disease Control and Prevention (CDC), spanning the years 2013-2023. The NHANES data are nationally representative, and the data collection process follows a multistage, stratified, and cluster design, in which in-house interviews with standardized examinations and laboratory tests are conducted through household-level mobile examination centers (MECs). The complex survey design yields population-level estimates that are reflective of the non-institutionalized U.S. population at the household level [9]. The study population consisted of 10,774 women aged 20-44 years, representing the reproductive age group. The data was filtered to include only the females who had been pregnant at least once. This yielded a sample size of 4,842 females across five cycles of the survey (2013-2023).

### Study objectives

The primary objective of this study was to evaluate the association of cardiometabolic and nutritional biomarkers with pregnancy loss among women of reproductive age in the United States. Specifically, the analysis sought to assess the following: (1) Estimate the associations between lipid biomarkers, Vitamin D levels, and Serum Folate Levels with pregnancy loss among women of reproductive age. (2) Quantify the association of socio-demographic and behavioral risk factors with pregnancy loss in the study population, and (3) evaluate the association of the composite nutritional risk score with pregnancy loss.

### Outcome variables

The primary endpoint was pregnancy loss. It was constructed in the following way using the available variables from the survey: Pregnancy Loss = Number of times pregnant – Number of Live Births. The pregnancy variable was then transformed into a dichotomous variable, with ‘Yes’ if pregnancy loss was ≥ 1 and ‘No’ indicating 0. It was then used to support the descriptive summary tables and logistic regression as the outcome variable.

### Exposure and covariate variables

#### Socio-demographic and behavioral covariates

The sociodemographic and Behavioral covariates included age, summarized as the Median with interquartile range (IQR), race/ethnicity, education level, smoking exposure, and BMI. Race/ ethnicity comprised of Mexican American, Non-Hispanic White, Non-Hispanic Black, Non-Hispanic Asian, and others composed of (Other Hispanic/ and other race/ multiracial). Education comprised of up to high school (as <9th grade; 9–11th (no diploma); High school/GED) as a single category and a second category of college and above (Some college/Associate’s; College graduate or above). For behavioral covariates, smoking exposure was constructed by labelling as “yes” if Serum cotinine was >10 ng/mL, and as “no” otherwise. BMI was categorized according to the WHO guidelines [10].

#### Laboratory and nutritional measures as covariates

All the laboratory measures of nutritional data were obtained from NHANES MEC standardized procedures. Nutritional variables were selected a priori based on prior literature linking them to pregnancy loss and were included according to data availability in NHANES, given substantial missingness across dietary measures. Lipid biomarkers were dichotomized as follows: Total Cholesterol (TC) >200 mg/dL (high), Low density Lipoprotein-cholesterol (LDL-C) >=160 mg/dL (High), High density Lipoprotein-Cholesterol (HDL-C) <60 mg/dL (Low), Triglycerides (TG) >=150 mg/dL (High) [11]. Deranged Lipid Profile variable was coded as “yes” if any of the TC, LDL-C, HDL-C, or TG met the abnormal thresholds; otherwise, it was coded as “no”. Serum Folate was categorised as Low if the value was < 10 nmol/L; otherwise, it was Considered Normal [12]. Vitamin D Levels were coded as insufficient if the value was 52.5 – 72.5 nmol/L and deficient if <52.5 nmol/L in accordance with the Institute of Medicine guidelines [13]. Composite Nutrition Risk Score of 0 to 3 was constructed to summarize cardiometabolic and micronutrient status, and categorized as follows: 0 if Normal Lipid and Nutrient Profile (Nutrient profile- Normal Folate and vitamin D levels), one if only lipid profile were deranged, two if only nutrient profile was deranged and lipid profile was typical, and three if both lipid and nutrient profile were deranged.

### Data preparation and bias control

Data integrity was maintained to ensure adherence to consistent units and categories. Primary descriptive analysis utilized complete-case estimations for each model, reflecting the actual sample size. No imputations were performed for the missing data.

### Statistical analysis

The analysis was performed in RStudio (version 2025.05.1+513). Boston, MA: RStudio, PBC; 2025) using several packages. Due to the complex survey design of NHANES, we incorporated MEC examination weights, strata, and primary sampling units’ weights wherever appropriate to ensure that the design-corrected estimates reflected the U.S. population. Non-parametric tests were used to test statistical significance. Descriptive variables were compared by pregnancy loss status, and the survey weighted methods (proportions) and the Chi-square test for categorical variables were used. The data were represented as counts and percentages wherever applicable. The multivariate logistic regression model reported adjusted odds ratios (aORs) with 95% confidence intervals, and *a P-value* <0.05 was denoted as statistically significant. The reference groups in the model used were: Non-Hispanic White for race/ethnicity, Normal class for BMI, College graduate or above for education, “No” for smoking exposure, and Normal for vitamin D, lipid, and micronutrient comparisons. Stepwise logistic regression was conducted to derive models for predicting pregnancy loss. Variables were removed or added using a combination of forward and backward elimination guided by the Akaike Information Criterion (AIC). Predictors with p<0.10 were retained. The bootstrapping procedure was then implemented to test the stability of predictors in the final model. Using the 2,000 resampled datasets, the frequency variable selection was recorded to identify stable predictors (≥70%), moderate predictors (40–70%), and unstable predictors (<40%) (Supplementary Table S1). The final multivariate logistic model included variables with stable and moderate values obtained from the bootstrapping method. Due to an inverse association of Vitamin D levels with pregnancy loss in model outcomes, we conducted a sensitivity analysis to evaluate the consistency of the effect estimates across different models involving different Vitamin D statuses (Model 1: without vitamin D, Model 2: with vitamin D, and Model 3: with collapsed Vitamin D levels (insufficient and deficient as ‘Low’)). We also examined the interaction of Vitamin D with lipid profile predictors and conducted an interaction analysis between vitamin D levels and Lipid Biomarkers (Supplementary Table S2) (Supplementary Table S3). Based on the results of these analyses, we removed the vitamin D variable from the final multivariate regression model due to its residual confounding effect and instability.

### Declaration of artificial intelligence (AI) tools

AI assistance (ChatGPT, OpenAI, San Francisco, CA, USA) was limited to generating some portions of the R code for statistical analysis. The code produced was manually reviewed, corrected, and validated by the authors prior to implementation to ensure its accuracy and appropriateness in accordance with the research objectives. The authors carefully reviewed the data, code and results in the manuscript for reproducibility and take full responsibility for the content of the publication. There was no other use of AI tools.

## Results

A total of 4,842 women aged 20-44 years with a history of at least one pregnancy were included from the NHANES dataset (2013-2023). The overall prevalence of pregnancy loss among U.S. women aged 20-44 years was 23% in the pooled 2013- 2023 NHANES dataset.

**Table 1** demonstrates that participants with a history of pregnancy loss were older than those without pregnancy loss (median age = 37.0 years [IQR 31.0–41.0] vs 35.0 years [IQR 30.0–40.0]; *P* -value < 0.001). Non-Hispanic Blacks had a higher prevalence of pregnancy loss compared with Non-Hispanic White women (25.8% vs 18.6%; *P* -value < 0.001). There were no significant differences by education level, though missingness was substantial across both groups. Participants with higher BMI (Class II, Class III) and smoking exposure had more pregnancy loss, and this difference was statistically significant.

**Table 1:** Distribution table for socio-demographic profile of study population pregnancy loss as outcome n, (%) (unadjusted)

| Variable | Pregnancy Loss |  | *P* -value <sup>b</sup> |
| --- | --- | --- | --- |
|  | Yes<br>(*n* =2510) | No<br>*n* =2332 |  |
| **Age in (years)**<br>**Median (IQR) <sup>a</sup>** | 37.0 (31.0-41.0) | 35.0 (30.0-40.0) | <0.001 |
| **Race/ Ethnicity** |  |  | <0.001 |
| Mexican American | 356 (14.2%) | 445 (19.1%) |  |
| Non-Hispanic Asian | 267 (10.6%) | 231 (9.9%) |  |
| Non-Hispanic Black | 648 (25.8%) | 434 (18.6%) |  |
| Non-Hispanic White | 787 (31.4%) | 866 (37.1%) |  |
| Other | 452 (18.0%) | 356 (15.3%) |  |
| **Education Level <sup>c</sup>** |  |  | 0.357 |
| Up to High School | 435 (17.3%) | 419 (18.0%) |  |
| College grad or above | 585 (23.3%) | 612 (26.2%) |  |
| Not Known/ Missing | 1490 (59.4%) | 1301 (55.8%) |  |
| **BMI (WHO)** |  |  | <0.001 |
| Normal (18.5–24.9) | 624 (24.9%) | 663 (28.4%) |  |
| Obesity I (30–34.9) | 463 (18.4%) | 447 (19.2%) |  |
| Obesity II (35–39.9) | 339 (13.5%) | 251 (10.8%) |  |
| Obesity III (≥40) | 381 (15.2%) | 279 (12.0%) |  |
| Overweight (25–29.9) | 653 (26.0%) | 617 (26.5%) |  |
| Underweight (<18.5) | 26 (1.0%) | 58 (2.5%) |  |
| Not Known/ Missing | 24 (1.0%) | 17 (0.7%) |  |
| **Smoking Exposure <sup>d</sup>** |  |  | <0.001 |
| No | 1866 (74.3%) | 1823 (78.2%) |  |
| Yes | 540 (21.5%) | 411 (17.6%) |  |
| Missing | 104 (4.1%) | 98 (4.2%) |  |

Values are *n* (%) unless otherwise specified. BMI = body mass index; WHO = World Health Organisation; IQR = interquartile range.
<sup>a</sup> Continuous variables are presented as median (IQR).
<sup>b</sup> *P*-values derived from the Wilcoxon rank-sum test (for age) and the $\chi^2$ test (for categorical variables).
<sup>c</sup> Missing data: Education = 55.8–59.4%
<sup>d</sup> Smoking exposure is defined as a serum cotinine level greater than 10 ng/mL.

**Table 2** demonstrates that higher odds of pregnancy loss were associated with an increase in each additional year in age (aOR = 1.02; 95% CI 1.00–1.04; *P*-value = 0.019). Compared with normal BMI, overweight (aOR = 1.56; 95% CI 1.16–2.11; p = 0.003), obesity I (aOR = 1.47; 95% CI 1.06–2.05; *P*-value = 0.022), and obesity II (aOR = 2.06; 95% CI 1.39–3.05; *P*-value < 0.001) were independently associated with increased risk. Women with smoking exposure had 1.58 times higher odds of pregnancy loss (95% CI 1.19–2.10; *P*-value = 0.002). Non-Hispanic Black ethnicity also remained significant (aOR = 1.32; 95% CI 1.00–1.74; *P* -value = 0.048). Educational attainment was not significantly associated after adjustment for other variables.

**Table 2.** Survey-weighted (MEC) multivariable logistic regression models for modifiable and non-modifiable risk factors (aOR, 95% CI)

| Variable | Model 1<br>(Non- Modifiable)<sup>a</sup> |  | Model 2<br>(Modifiable)<sup>b</sup> |  |
| --- | --- | --- | --- | --- |
|  | aOR (95% CI) | *P* -value<sup>c</sup> | aOR (95% CI) | *P* -value<sup>c</sup> |
| **Age** |  |  |  |  |
| (per 1 unit) | 1.02 (1.00-1.04) | 0.013 | 1.02 (1.00 - 1.04) | 0.019 |
| **Race/ Ethnicity** |  |  |  |  |
| Non-Hispanic White (ref) | 1 |  | 1 |  |
| Mexican American | 0.81 (0.62- 1.06) | 0.127 | 0.81 (0.61- 1.08) | 0.155 |
| Non-Hispanic Asian | 0.86 (0.62 -1.18) | 0.349 | 1.08 (0.76- 1.55) | 0.654 |
| Non-Hispanic Black | 1.48 (1.14- 1.91) | 0.003 | 1.32 (1.00- 1.74) | 0.048 |
| Other (multi-racial) | 1.17 (0.87- 1.57) | 0.307 | 1.15(0.85- 1.56) | 0.374 |
| **Education** |  |  |  |  |
| College grad or above (ref) | 1 |  | 1 |  |
| Up to High School | 1.25 (1.00- 1.56) | 0.049 | 1.08 (0.85- 1.36) | 0.533 |
| **BMI (WHO)** |  |  |  |  |
| Normal (18.5,24.9) (ref) | 1 |  | 1 |  |
| Obesity I (30,34.9) |  |  | 1.47 (1.06- 2.05) | 0.022 |
| Obesity II (35,39.9) |  |  | 2.06 (1.39- 3.05) | <0.001 |
| Obesity III (>40) |  |  | 1.48 (1.01- 2.17) | 0.047 |
| Overweight (25,29.9) |  |  | 1.56 (1.16- 2.11) | 0.003 |
| Underweight (<18.5) |  |  | 0.49- 2.76 | 0.737 |
| **Smoking exposure** |  |  |  |  |
| No (ref) | 1 |  | 1 |  |
| Yes |  |  | 1.58 (1.19- 2.10) | 0.002 |

aOR = adjusted odds ratio; CI = confidence interval; ref = reference category. Survey weights, strata, and PSU design variables are applied to account for the complex sampling design of NHANES.
<sup>a</sup> Model 1 = non-modifiable variables (age, race, education).
<sup>b</sup> Model 2 = Model 1 + modifiable factors (BMI, smoking exposure).
<sup>c</sup> *P*-values are two-sided and reflect Wald tests.

**Table 3** shows that women with a history of pregnancy loss had significantly higher levels of total cholesterol, LDL-C, and triglycerides, and lower levels of HDL-C, compared with those without loss (all p < 0.05). A deranged lipid profile was observed in 22% of women compared to 19% without pregnancy loss (*P*-value = 0.024). No significant group differences were observed in serum folate or vitamin D concentrations. Although a slightly higher proportion of vitamin D deficiency (37% vs 36%) was seen among women with pregnancy loss, the association did not reach statistical significance (*P*-value = 0.058). When integrating both micronutrient and lipid abnormalities into a composite nutritional risk score, higher scores (≥2) were associated with an increased frequency of pregnancy loss (*P*-value = 0.026).

**Table 3.**
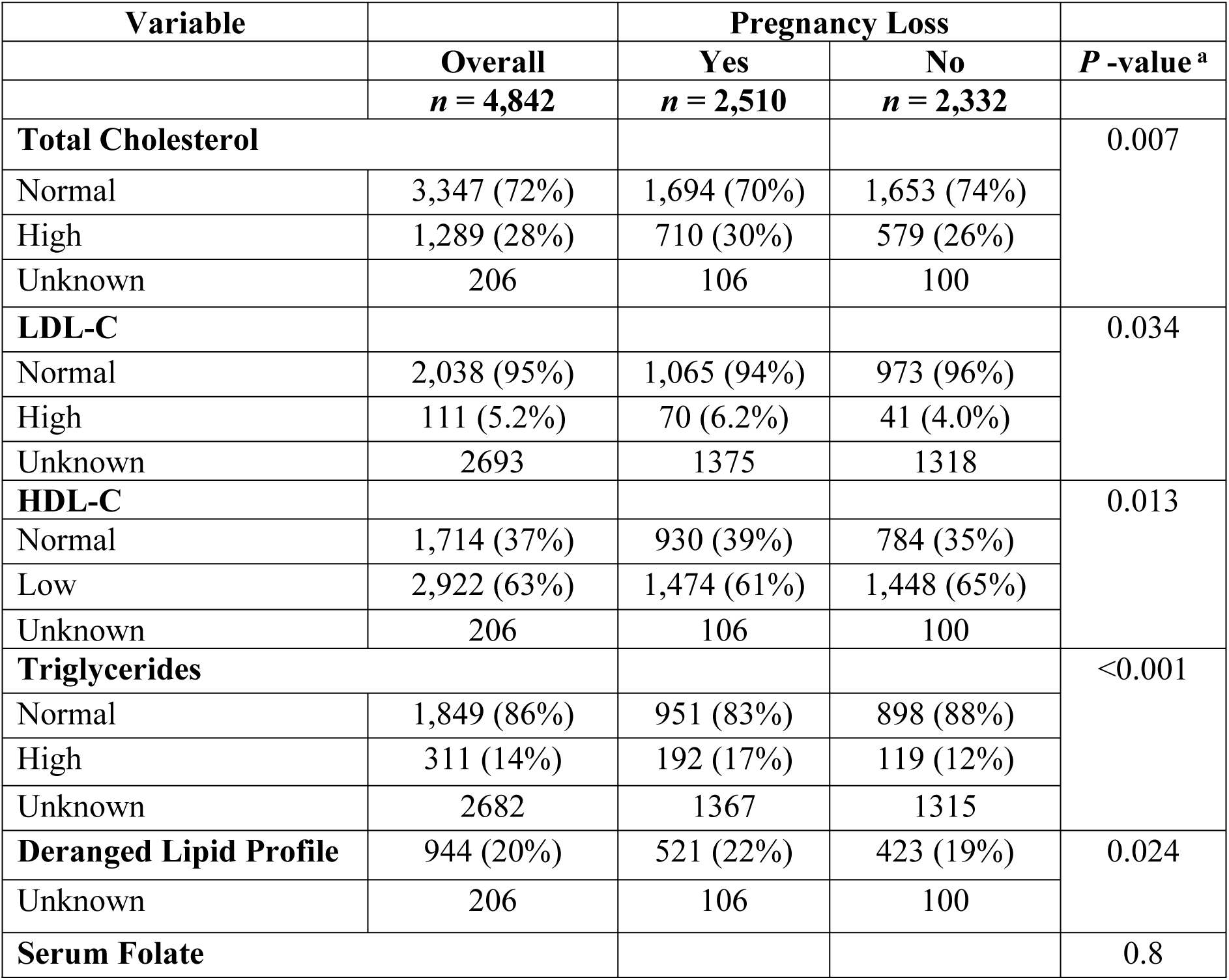
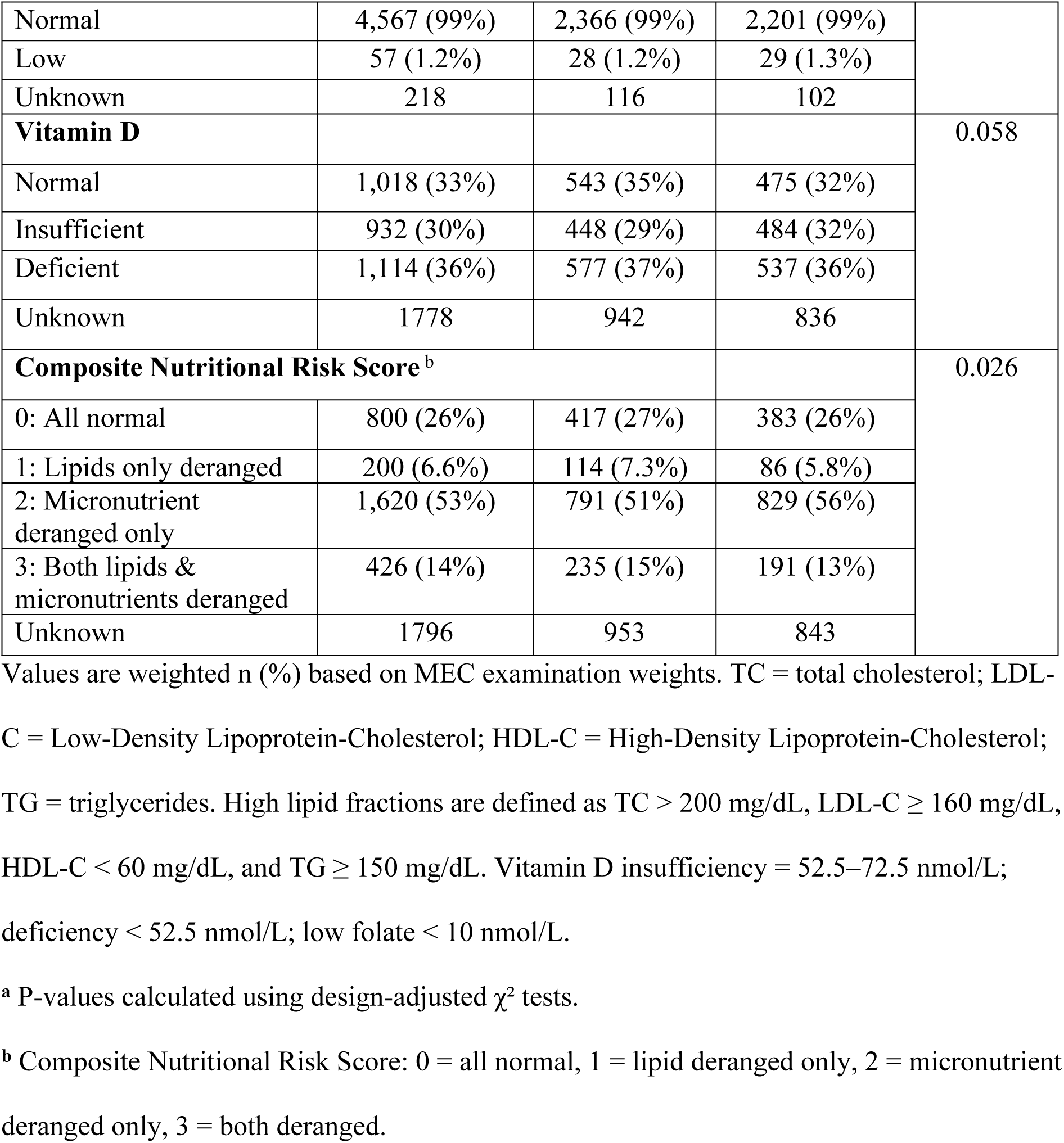
Distribution table for nutritional profile of study population for pregnancy loss among study population n, (%)

Sensitivity models incorporating vitamin D categories did not alter the overall associations. Although low vitamin D showed a protective direction of effect (aOR = 0.67; 95% CI 0.05–0.94; *P*-value = 0.019), the result was deemed biologically implausible and excluded from the final regression models (**Table 4**). Moreover, Bootstrapping analysis, Interaction models between vitamin D levels and lipid biomarkers deemed vitamin D as an unstable factor and generated a protective effect on pregnancy loss likely due to residual confounding and sparse sample size within interaction groups (**S1-S3 Table**).

**Table 4:** Sensitivity analyses models for vitamin D.

| Variables | Model 1<br>(without Vitamin D) <sup>a</sup> |  | Model 2<br>(with Vitamin D) <sup>b</sup> |  | Model 3<br>(Collapsed Vitamin D categories) <sup>c</sup> |  |
| --- | --- | --- | --- | --- | --- | --- |
|  | Estimate<br>(95% CI) <sup>d</sup> | P-value | Estimate<br>(95% CI)<sup>d</sup> | P-value | Estimate<br>(95% CI)<sup>d</sup> | P-value |
| (Intercept) | 0.855<br>(0.51 - 1.43) | 0.552 | 0.921<br>(0.554- 1.53) | 0.75 | 0.91<br>(0.55 -1.51) | 0.716 |
| **Race/ Ethnicity** |  |  |  |  |  |  |
| Non-Hispanic Asian | 0.976<br>(0.56- 1.68) | 0.929 | 0.962<br>(0.556- 1.66) | 0.888 | 0.971<br>(0.56 -1.68) | 0.917 |
| Non-Hispanic Black | 1.82<br>(1.11 -2.97) | 0.016 | 1.92<br>(1.17- 3.16) | 0.009 | 1.86<br>(1.14 -3.04) | 0.013 |
| Non-Hispanic White | 0.765<br>(0.49 -1.18) | 0.222 | 0.733<br>(0.47- 1.13) | 0.16 | 0.758<br>(0.49 -1.16) | 0.206 |
| Other | 1.21<br>(0.73 -2) | 0.468 | 1.21<br>(0.73- 2) | 0.467 | 1.22<br>(0.74 -2.03) | 0.432 |
| **BMI (WHO)** |  |  |  |  |  |  |
| Obesity I<br>(30–34.9) | 1.44<br>(0.92 -2.27) | 0.11 | 1.43<br>(0.91 -2.24) | 0.122 | 1.42<br>(0.91 -2.23) | 0.126 |
| Obesity II<br>(35–39.9) | 2.12<br>(1.23 -3.65) | 0.006 | 2.06<br>(1.21 -3.55) | 0.008 | 2.07<br>(1.21 -3.56) | 0.008 |
| Obesity III (≥40) | 1.86<br>(1.06 -3.28) | 0.03 | 1.86<br>(1.06- 3.28) | 0.030 | 1.82<br>(1.04 -3.2) | 0.036 |
| Overweight<br>(25–29.9) | 1.68<br>(1.12 -2.52) | 0.012 | 1.7<br>(1.13- 2.55) | 0.010 | 1.69<br>(1.13 -2.54) | 0.010 |
| Underweight (<18.5) | 0.36<br>(0.08 -1.24) | 0.135 | 0.367<br>(0.079- 1.27) | 0.143 | 0.367<br>(0.08 -1.26) | 0.142 |
| **Smoking exposure**<br>**(Yes)** | 1.33<br>(0.91 -1.95) | 0.141 | 1.35<br>(0.926- 1.99) | 0.119 | 1.33<br>(0.91 -1.95) | 0.138 |
| **LDL-C (High)** | 3.66<br>(1.53 -9.34) | 0.004 | 2.93<br>(1.42- 6.51) | 0.005 | 2.97<br>(1.45 -6.61) | 0.004 |
| **HDL-C (Low)** | 0.721<br>(0.51- 1.01) | 0.058 | 0.697<br>(0.5- 0.97) | 0.032 | 0.695<br>(0.50 -0.97) | 0.031 |
| **TG (High)** | 2.34<br>(1.2 -4.68) | 0.013 | 1.86<br>(1.18- 2.96) | 0.008 | 1.83<br>(1.16 -2.9) | 0.009 |
| **Vitamin D** |  |  |  |  |  |  |
| Insufficient |  |  | 0.732<br>(0.50- 1.06) | 0.101 |  |  |
| Deficient |  |  | 0.606<br>(0.41- 0.89) | 0.011 |  |  |
| Vitamin D Low<br>(insufficient<br>+Deficient) |  |  |  |  | 0.671<br>(0.05 -0.94) | 0.019 |

BMI = Basal Metabolic Index; HDL-C = High Density Lipoprotein-Cholesterol; LDL-C = Low Density Lipoprotein-Cholesterol; TG = Triglycerides; WHO = World Health Organisation; CI = Confidence Interval. Models 1–3 progressively incorporate vitamin D variables
<sup>a</sup> Model 1: without vitamin D
<sup>b</sup> Model 2: with vitamin D categories insufficient and deficient
<sup>c</sup> Model 3: with collapsed Vitamin D categories of insufficient and deficient.
<sup>d</sup> Estimate = adjusted odds ratio; CI = confidence interval.

Bootstrapping analyses demonstrated high stability (≥ 70%) for the following categories: race (Non-Hispanic Black), HDL-C (low), triglycerides (high), and BMI (Underweight, Obesity II–III), confirming the model’s robustness (**S1 Table**).

**Fig 1**, illustrates that the distribution of pregnancy loss varies across composite nutritional risk categories (*P*-value = 0.002). In the pairwise chi-square comparison, women with only micronutrient derangements had the highest proportion of pregnancy loss, and this difference was statistically significant from other groups. This suggests that micronutrient abnormalities alone may contribute to the risk of pregnancy loss than lipid abnormalities alone. This difference can also be attributed to the difference in the frequency composition of the micronutrient derangement group when compared to other categories. Higher composite nutritional risk scores were associated with increased risk.

**Fig 1.**
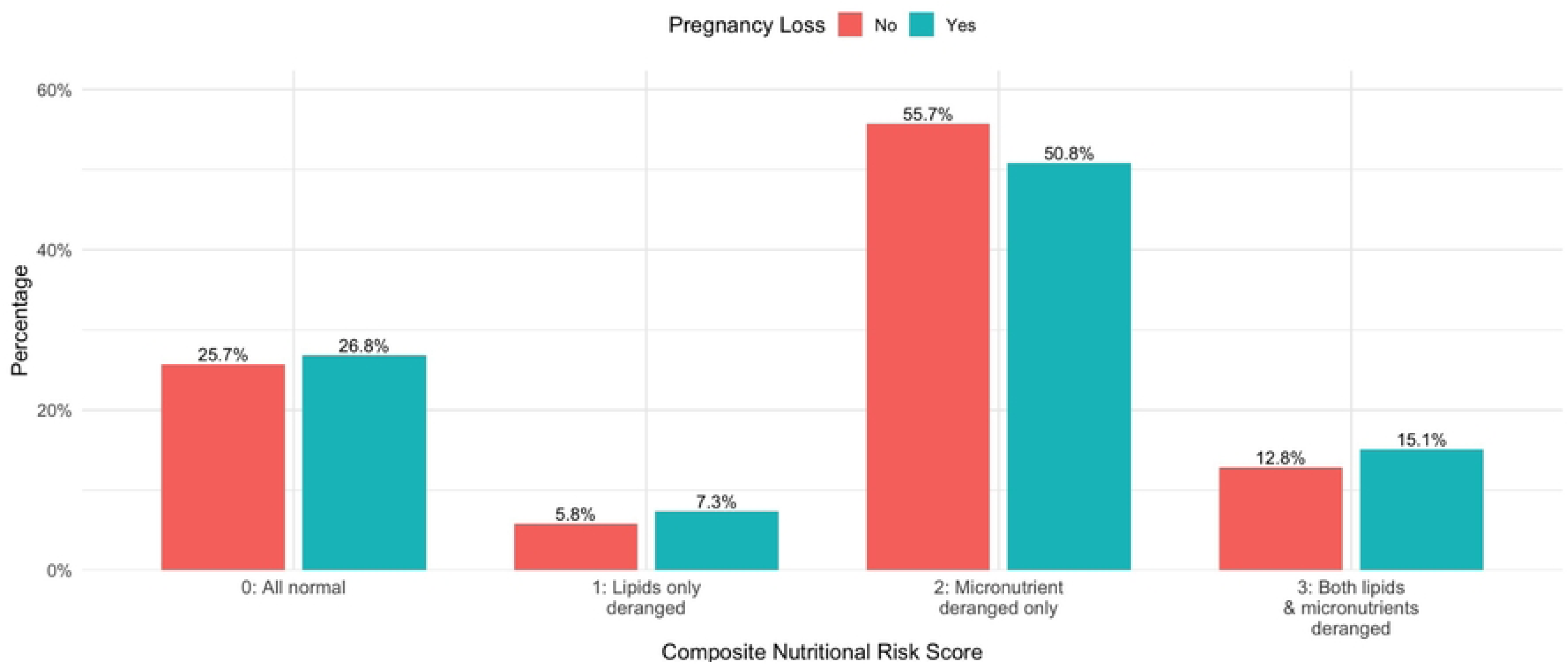
Composite nutritional risk Score by pregnancy loss (%).

**Fig 2**, illustrates adjusted odds ratios for predictors of pregnancy loss in the final multivariable model. Obesity class II (aOR = 1.79; 95% CI 1.28–2.51; *P*-value = 0.0007) and class III (aOR = 1.68; 95% CI 1.21–2.33; *P*-value = 0.0019) were strongly associated with higher odds of pregnancy loss, whereas underweight women had significantly lower odds (aOR = 0.13; 95% CI 0.04–0.31; *P*-value < 0.001). Low HDL-C (aOR = 0.68; 95% CI 0.55–0.83; *P*-value = 0.0002) and high triglycerides (aOR = 1.89; 95% CI 1.44–2.48; *P*-value < 0.001) were also independent predictors. Among racial groups, Non-Hispanic Black (aOR = 2.00; 95% CI, 1.49–2.69; *P*-value < 0.001) and the “Other” race (aOR = 1.68; 95% CI, 1.23–2.30; *P*-value = 0.0011) were associated with an increased risk. No significant associations were observed for LDL-C, smoking exposure, or overweight BMI.

**Fig 2.**
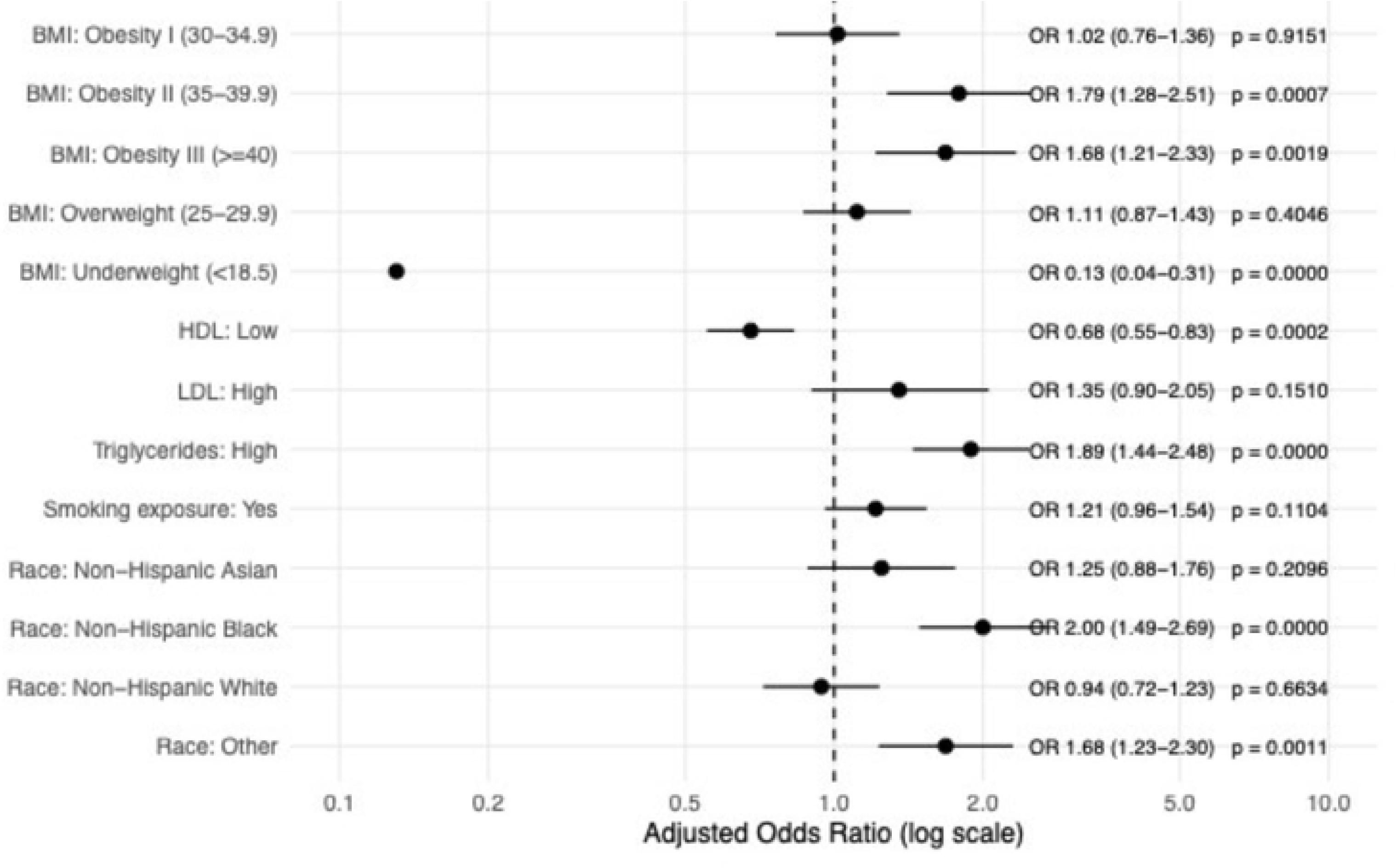
Forest plot depicting the Multivariate Regression for Pregnancy Loss.

## Discussion

This study provides population-level evidence on the association between routinely measured cardiometabolic and behavioral markers and a history of pregnancy loss among U.S. women of reproductive age. Using nationally representative NHANES data and survey-weighted analyses, we identified a consistent pattern in which higher BMI, adverse lipid profiles, and smoking exposure clustered among women reporting prior pregnancy loss. These findings are best interpreted as identifying risk markers rather than causal determinants, given the cross-sectional design. Consistent with national estimates, approximately one in four reproductive-age women reported experiencing a pregnancy loss during this period, highlighting the substantial population-level burden of this outcome [2]. Importantly, elevated LDL-C and triglyceride levels were associated with nearly double the odds of pregnancy loss. These findings support a multifactorial etiology for pregnancy loss, emphasizing modifiable behavioral and metabolic factors as critical contributors.

### Maternal age, race and lifestyle factors

Advancing maternal age was independently associated with higher odds of pregnancy loss, consistent with established evidence linking increasing age to diminished oocyte quality, chromosomal abnormalities, and reduced uterine receptivity [14]. The higher odds observed among Non-Hispanic Black women align with prior evidence on persistent racial disparities in adverse reproductive outcomes in the U.S., likely influenced by differences in healthcare access, chronic stress, and socioeconomic inequities [15]. Higher BMI and smoking exposure were also confirmed as risk factors for pregnancy loss, consistent with prior literature [16]. Obesity may disrupt pregnancy through systemic inflammation and impaired placental function [7] and is often accompanied by dyslipidaemia or a deranged lipid profile.

### Cardiometabolic risk: HDL-C, LDL-C and triglycerides

Elevated triglycerides and low HDL-C emerged as the most stable lipid-related predictors across multivariable and bootstrap analyses. Potential mechanisms include placental inflammation and vascular dysregulation [17]. Metabolomic studies further suggest that disruptions in fatty acid and folate pathways may underlie miscarriage, highlighting potential biomarker avenues for earlier prognosis [18]. While the biological mechanisms linking dyslipidaemia to pregnancy loss are not fully understood, lipid abnormalities may reflect broader metabolic dysfunction relevant to early pregnancy. From a clinical epidemiology perspective, lipid biomarkers may serve as accessible indicators of metabolic risk, rather than direct causal agents, particularly in women with obesity, genetic predisposition or other cardiometabolic conditions or in settings where detailed metabolic phenotyping is not feasible.

### Nutritional factors: vitamin D and folate

Low vitamin D levels disrupt maternal immune tolerance and trophoblastic invasion, processes critical for maintaining gestation [19], and prior studies have linked vitamin D deficiency to an increased risk of miscarriage [19] [5]. Similarly, folate is essential for DNA synthesis and embryonic development [20] [21]. However, Vitamin D and folate were not consistently associated with pregnancy loss after accounting for confounding and predictor instability. Sensitivity and interaction analyses suggested that vitamin D estimates were sensitive to model specification and sample composition, leading to biologically implausible effect directions.

Further, widespread folate fortification and supplementation in the U.S., likely limits variability in NHANES data. Additionally, higher composite nutritional risk scores were associated with increased risk for pregnancy loss highlighting the role of multiple nutritional deficiencies. These findings highlight the challenges of interpreting single time-point micronutrient measurements in cross-sectional surveys and underscore the importance of cautious interpretation in observational nutritional epidemiology.

### Clinical and epidemiologic implications

From a clinical epidemiology standpoint, the observed associations suggest that pregnancy loss may cluster with established cardiometabolic risk profiles already used in routine care. While causal inference cannot be established, these findings support further evaluation of cardiometabolic markers as part of risk stratification frameworks in preconception and inter-conception settings. Longitudinal studies are needed to determine whether modification of these risk markers alters subsequent pregnancy outcomes.

### Strengths and limitations

Key strengths include the use of a nationally representative dataset, standardized biomarker measurement, and formal assessment of predictor stability using bootstrap resampling. Limitations include the cross-sectional design which precludes causal inference, reliance on self-reported reproductive history, which may introduce recall bias and inadequate laboratory measurements, and lack of temporal alignment between biomarker measurement and pregnancy loss events. Additionally, given the inability to ascertain the timing of nutritional assessments relative to pregnancy and incomplete availability of some dietary variables, some associations may also have been underestimated because of non-differential exposure misclassification.

## Conclusions

This study identifies a consistent association between pregnancy loss and modifiable cardiometabolic and behavioral risk markers in a nationally representative U.S. sample. Our results do not establish causality but are intended to inform risk assessment and highlight the potential role of metabolic risk profiling in identifying women at higher risk. Future prospective studies are needed to clarify temporal relationships and inform evidence-based strategies for prevention and counselling. These findings reinforce the need for targeted interventions addressing modifiable cardiometabolic and lifestyle risk factors with a particular emphasis on monitoring and managing dyslipidaemia, to improve pregnancy outcomes and long-term maternal health, which is not routine in clinical practice during pregnancy. Future research should aim to explore careful precision nutritional and metabolic assessments to establish causal relationships and identify high-risk individuals early and guide multidisciplinary prevention strategies.

## Data Availability

The datasets were derived from publicly available sources. The data for this research can be accessed through the CDC NHANES website by accessing the following URL: https://www.cdc.gov/nchs/nhanes/index.html. We are committed to data sharing that we downloaded, cleaned, and cleaned to generate secondary variables for use in the analysis, and any requests for data or code access should be made to the corresponding author

## Supporting information

**S1 Table. Bootstrapping model to test model stability**. BMI= Basal Metabolic Index; HDL= High Density Lipoprotein-Cholesterol; LDL= Low Density Lipoprotein-Cholesterol. Selection frequency = percentage of 2000 bootstrap samples in which the variable was retained. ≥ 70% = stable; 40–70% = moderate; < 40% = unstable. Variables with high selection frequency indicate robust predictors across resampling iterations.

**S2 Table. Interaction models summary between vitamin D and lipid abnormality.** BMI= Basal Metabolic Index; HDL-C = High Density Lipoprotein-Cholesterol; LDL-C = Low Density Lipoprotein-Cholesterol; TGs= Triglycerides; df= degree of freedom; χ^2^= Chi Square estimate; aOR= Adjusted odd’s ratio; CI= Confidence Interval.

**S3 Table. Stratified analysis based on interaction models.** HDL-C = High Density Lipoprotein-Cholesterol; TGs= Triglycerides; aOR= Adjusted odd’s ratio; CI= Confidence Interval

## References

1. ACOG Practice Bulletin No. 200: Early Pregnancy Loss. Obstet Gynecol. 2018;132(5):e197–e207. doi: 10.1097/aog.0000000000002899. PubMed PMID: 30157093.

2. Forrest SE, Rossen LM, Ahrens KA. Trends in Risk of Pregnancy Loss Among US Women by Metropolitan Status, 2000-2018. Paediatr Perinat Epidemiol. 2025. Epub 20250927. doi: 10.1111/ppe.70066. PubMed PMID: 41014169; PubMed Central PMCID: PMCPMC12477614.

3. Boxem AJ, Blaauwendraad SM, Mulders A, Bekkers EL, Kruithof CJ, Steegers EAP, et al. Preconception and Early-Pregnancy Body Mass Index in Women and Men, Time to Pregnancy, and Risk of Miscarriage. JAMA Netw Open. 2024;7(9):e2436157. Epub 20240903. doi: 10.1001/jamanetworkopen.2024.36157. PubMed PMID: 39298166; PubMed Central PMCID: PMCPMC11413718.

4. Pineles BL, Park E, Samet JM. Systematic review and meta-analysis of miscarriage and maternal exposure to tobacco smoke during pregnancy. Am J Epidemiol. 2014;179(7):807–23. Epub 20140210. doi: 10.1093/aje/kwt334. PubMed PMID: 24518810; PubMed Central PMCID: PMCPMC3969532.

5. Tamblyn JA, Pilarski NSP, Markland AD, Marson EJ, Devall A, Hewison M, et al. Vitamin D and miscarriage: a systematic review and meta-analysis. Fertility and Sterility. 2022;118(1):111–22. doi: 10.1016/j.fertnstert.2022.04.017.

6. De-Regil LM, Peña-Rosas JP, Fernández-Gaxiola AC, Rayco-Solon P. Effects and safety of periconceptional oral folate supplementation for preventing birth defects. Cochrane Database of Systematic Reviews. 2015;(12). doi: 10.1002/14651858.CD007950.pub3. PubMed PMID: CD007950.

7. Preda A, Preda SD, Mota M, Iliescu DG, Zorila LG, Comanescu AC, et al. Dyslipidemia in Pregnancy: A Systematic Review of Molecular Alterations and Clinical Implications. Biomedicines. 2024;12(10). Epub 20241003. doi: 10.3390/biomedicines12102252. PubMed PMID: 39457565; PubMed Central PMCID: PMCPMC11504282.

8. Chen TC, Clark J, Riddles MK, Mohadjer LK, Fakhouri THI. National Health and Nutrition Examination Survey, 2015-2018: Sample Design and Estimation Procedures. Vital Health Stat 2. 2020;(184):1–35. PubMed PMID: 33663649.

9. National Health and Nutrition Examination Survey (NHANES) [Internet]. 2024. Available from: https://www.cdc.gov/nchs/nhanes/index.html.

10. Adult BMI categories [Internet]. 2024. Available from: https://www.cdc.gov/bmi/adult-calculator/bmi-categories.html.

11. Yang HS. Lipid Biomarkers and Cardiometabolic Diseases: Critical Knowledge Gaps and Future Research Directions. Metabolites. 2025;15(2). Epub 20250207. doi: 10.3390/metabo15020108. PubMed PMID: 39997733; PubMed Central PMCID: PMCPMC11857555.

12. Mamme NY, Roba HS, Fite MB, Asefa G, Abrahim J, Yuya M, et al. Serum folate deficiency and associated factors among pregnant women in Haramaya District, Eastern Ethiopia: a community-based study. BMJ Open. 2023;13(5):e068076. Epub 20230508. doi: 10.1136/bmjopen-2022-068076. PubMed PMID: 37156586; PubMed Central PMCID: PMCPMC10174039.

13. Rosen CJ, Abrams SA, Aloia JF, Brannon PM, Clinton SK, Durazo-Arvizu RA, et al. IOM committee members respond to Endocrine Society vitamin D guideline. J Clin Endocrinol Metab. 2012;97(4):1146–52. Epub 20120322. doi: 10.1210/jc.2011-2218. PubMed PMID: 22442278; PubMed Central PMCID: PMCPMC5393439.

14. Glick I, Kadish E, Rottenstreich M. Management of Pregnancy in Women of Advanced Maternal Age: Improving Outcomes for Mother and Baby. International Journal of Women’s Health. 2021;13(null):751–9. doi: 10.2147/IJWH.S283216.

15. Saluja B, Bryant Z. How Implicit Bias Contributes to Racial Disparities in Maternal Morbidity and Mortality in the United States. J Womens Health (Larchmt). 2021;30(2):270–3. Epub 20201125. doi: 10.1089/jwh.2020.8874. PubMed PMID: 33237843.

16. Mirakbarova Z, Pascat V, Akramkhanova S, Chu C-Y, Yusupov U, Scapoli C, et al. Genetics of Recurrent Miscarriage and Pregnancy Loss in Women’s Reproductive Health. medRxiv. 2025:2025.02.15.25321247. doi: 10.1101/2025.02.15.25321247.

17. Chen L, Mei-Ling Tan K, Leow MK-S, Tan KH, Yen Chan JK, Chan S-Y, et al. Characterisation of pregnancy-induced alterations in apolipoproteins and their associations with maternal metabolic risk factors and offspring birth outcomes: a preconception and longitudinal cohort study. eBioMedicine. 2025;112:105562. doi: 10.1016/j.ebiom.2025.105562.

18. Ku CW, Tan YB, Chew KY, Ku CO, Ng STC, Tan TC, et al. Untargeted metabolomics reveals key pathways in miscarriage: steroid, folate, fatty acid & glycosaminoglycan metabolism. npj Women’s Health. 2025;3(1):35. doi: 10.1038/s44294-025-00085-9.

19. Chien M-C, Huang C-Y, Wang J-H, Shih C-L, Wu P. Effects of vitamin D in pregnancy on maternal and offspring health-related outcomes: An umbrella review of systematic review and meta-analyses. Nutrition & Diabetes. 2024;14(1):35. doi: 10.1038/s41387-024-00296-0.

20. Lei F, Zhang L, Wang L, Wu W, Wang F. Association between early spontaneous abortion and homocysteine metabolism. Frontiers in Medicine. 2024;Volume 11 - 2024. doi: 10.3389/fmed.2024.1310112.

21. Gaskins AJ, Rich-Edwards JW, Hauser R, Williams PL, Gillman MW, Ginsburg ES, et al. Maternal prepregnancy folate intake and risk of spontaneous abortion and stillbirth. Obstet Gynecol. 2014;124(1):23–31. doi: 10.1097/aog.0000000000000343. PubMed PMID: 24901281; PubMed Central PMCID: PMCPMC4086728.

